# Artificial intelligence-based automated left ventricular mass quantification from non-contrast cardiac CT scans: correlation with contrast CT and cardiac MRI

**DOI:** 10.1101/2024.01.12.24301169

**Authors:** Donghee Han, Aakash Shanbhag, Robert JH Miller, Nicholas Kwok, Parker Waechter, Valerie Builoff, David E Newby, Damini Dey, Daniel S Berman, Piotr Slomka

**Author notes:** Authors contributed equally (Donghee Han, Aakash Shanbhag). Corresponding Author: Piotr Slomka, PhD, FACC Cedars-Sinai Medical Center 6500 Wilshire Blvd, Los Angeles, California 90048.

## Abstract

**Background:** Non-contrast CT scans are not used for evaluating left ventricle myocardial mass (LV mass), which is typically evaluated with contrast CT or cardiovascular magnetic resonance imaging (MRI). We assessed the feasibility of LV mass estimation from standard, ECG-gated, non-contrast CT using an artificial intelligence (AI) approach and compare it with coronary CT angiography (CTA) and cardiac MRI.

**Methods:** We enrolled consecutive patients who underwent coronary CTA, which included non-contrast CT calcium scanning and contrast CTA, and cardiac MRI. The median interval between coronary CTA and MRI was 22 days (IQR: 3-76). We utilized an nn-Unet AI model that automatically segmented non-contrast CT structures. AI measurement of LV mass was compared to contrast CTA and MRI.

**Results:** A total of 316 patients (Age: 57.1±16.7, 56% male) were included. The AI segmentation took on average 22 seconds per case. An excellent correlation was observed between AI and contrast CTA LV mass measures (r=0.84, p<0.001), with no significant differences (136.5±55.3 vs. 139.6±56.9 g, p=0.133). Bland-Altman analysis showed minimal bias of 2.9. When compared to MRI, measured LV mass was higher with AI (136.5±55.3 vs. 127.1±53.1 g, p<0.001). There was an excellent correlation between AI and MRI (r=0.85, p<0.001), with a small bias (−9.4). There were no statistical differences between the correlations of LV mass between contrast CTA and MRI, or AI and MRI.

**Conclusions:** The AI-based automated estimation of LV mass from non-contrast CT demonstrated excellent correlations and minimal biases when compared to contrast CTA and MRI.

## Introduction

Accurate assessment of left ventricle myocardial mass (LV mass) is crucial for diagnosis and prognostication of various cardiac diseases ^1–4^. While echocardiography and cardiac magnetic resonance imaging (MRI) are commonly used for evaluating cardiac structure, contrast cardiac CT can also provide cardiac structure information ^5–7^. With its high spatial resolution and contrast-to-noise signal with contrast administration, contrast-enhanced cardiac CT angiography (CTA) has demonstrated accurate assessment of cardiac chambers and functions ^6^.

Non-contrast coronary artery calcium (CAC) CT scanning is a widely used and rapidly growing procedure for assessing the burden of coronary atherosclerosis ^8–10^. However, it has not been employed to date for cardiac structural evaluation due to the absence of contrast enhancement. In particular, non-contrast CT has not been employed to date for evaluating LV mass, which is a well-established prognostic indicator for cardiovascular risk ^11,12^.

Non-contrast CT scans exhibit similar CT attenuation between the LV myocardium and blood pool, making it challenging to measure LV mass ^13^. Recent advancements in artificial intelligence (AI) and deep learning have shown promise in enhancing the identification of inferred anatomy from learned examples that may not be detectable through routine visual evaluation of non-contrast CT alone ^14,15^. In this study, we aimed to assess the feasibility of LV mass estimation from standard non-contrast CAC scans using an AI approach and compare it with clinically reported LV mass from contrast CT and MRI.

## Methods

### Study population

We retrospectively assessed consecutive patients who underwent coronary CTA, which included non-contrast CAC scanning and contrast-enhanced CTA, and cardiac MRI within a 12-month interscan interval at Cedars-Sinai Medical Center from January 2015 to May 2022. The study protocol complied with the Declaration of Helsinki and was approved by the institutional review board. Written informed consent was waived by the institutional review board due to the retrospective nature of the study. To the extent allowed by data sharing agreements and institutional review board protocols, the data and code from this manuscript will be shared upon written request.

### Clinical data

At the time of CTA or MRI scanning, all patients completed a questionnaire regarding clinical symptoms, cardiovascular risk factors, current medications, and previous cardiac interventions. Premature familial coronary artery disease (CAD) history was defined as a primary relative diagnosed for CAD or cardiac event <55 years for male family member or <65 for female family member. Smoking was defined as either currently smoking or having stopped for <1 year. Hypertension, hypercholesterolemia, and diabetes were defined based on self-reported history.

### CTA acquisition and LV mass evaluation

CTA images, including both non-contrast CAC scanning and contract enhanced coronary CTA, were acquired using dual-source CT scanners (Somatom Flash, or Force, Siemens Healthineers). Contrast-enhanced coronary CTA images were acquired using the following protocol: Prospective electrocardiogram (ECG) gating at either end systole or mid diastole or helical with dose modulation, with tube voltage (80-120 kVp), and current 300–700 mA, and the tube voltage and current were adjusted by experienced radiologic technologists. In preparation for image acquisition, patients were administered 90–120 mL of intravenous contrast (Omnipaque or Visipaque, GE Healthcare, Princeton, New Jersey, USA) at a rate of 4–7 mL/s, sublingual nitrates, and intravenous or oral beta-blockers if indicated for pre-scan heart rate reduction. Images were reconstructed with a slice thickness of 0.6 mm, an increment of 0.5 mm, 250-mm field of view, and 512 × 512 matrix.

The LV mass was measured by experienced cardiologists from contrast-enhanced coronary CTA using a dedicated workstation (Syngo.via, Siemens Healthineers) based on end-systolic or mid-diastolic phase during standard clinical review. The workstation automatically detected the endocardial and epicardial myocardial borders, which were then manually edited by the readers as required. The LV mass volume excluded the papillary muscle.

### MRI acquisition and LV mass evaluation

All MRI scans were performed using a 1.5T scanner (Avanto, Siemens Healthineers, Erlangen, Germany). Retrospectively gated cine steady-state free precession images were obtained in multiple planes, including a stack of short-axis slices with coverage from the LV base to the apex (6 mm slice thickness, 2-mm interslice gap).

To assess LV mass, LV endocardial and epicardial borders at end-diastole were manually contoured on short-axis steady-state free precession images by experienced cardiologists using a workstation (Vitrea, Vital Images Inc., Minnetonka, Minnesota). Papillary muscles were excluded from LV mass. All measurements for CTA and MRI were retrieved from the clinical database.

### Non-contrast CT acquisition

Non-contrast CT images were acquired as a part of the coronary CTA for CAC assessment, using a scan protocol of prospective ECG triggering at 50–80% of the cardiac cycle, with 3 mm slice thickness, 120-kVp tube voltage, automated tube current adjustment based on patient body size. All non-contrast CT was reconstructed into matrix size of 512×512, 3-mm slice thickness, and using the standard B35f kernel.

### AI myocardial segmentation

LV myocardium was segmented from gated non-contrast CT scans, which were acquired for CAC scoring during CTA scanning, using Total Segmentator (TS) model developed by Wasserthal et al ^16^. TS utilizes a no new-net UNet (nnU-Net) architecture to automatically segment a variety of anatomic structures from images ^17^. The TS model was previously trained using expert annotations from contrast images which were transferred to registered non-contrast images, on a subset of the 1204 CT examinations. A standard density factor of 1.055 g/ml was used to convert LV myocardial volume to LV mass calculation.

### Statistical analysis

Data are expressed as mean ± standard deviation for continuous variables. Categorical variables are presented as a number (percentage). Data were analyzed by paired two-sided t-tests or Wilcoxon matched-pairs signed-rank test as appropriate. Bland Altman analysis was carried out to compare the LV mass between AI, contrast CTA, and MRI, and used to estimate the bias and 95% confidence intervals. Spearman’s rank order correlation was used to compare LV mass between AI, and contrast CTA, and MRI. Additional subgroup analyses were also performed on the bases of sex and for the patients who were referred for cardiomyopathy evaluation. A two-sided p-value of less than 0.05 was considered statistically significant. Python (version 3.74) and libraries scikit learn stats^18^and pycompare^19^ were used to estimate these metrics. Significance comparison for p values was carried out on STATA (version 17; StataCorp, College Station, TX, USA).

## Results

### Baseline characteristics

Baseline clinical characteristics of the study population are shown in **Table 1**. The mean age was 57.1±16.7 and 56% were male. Participants had a mean body mass index of 26.7±5.6. Approximately half of the study population had a history of hypertension (50.3%) or hypercholesterolemia (45.9%). The proportion of patients with prior CAD history was 13.9%.

**Table 1.** Baseline Characteristics of the study population.

| **Number of patients (n=316)** |  |
| --- | --- |
|  | **Values** |
| **Age** | 57.1±16.7 |
| **Male sex** | 177 (56.0) |
| **Body mass index** | 26.7±5.6 |
| **Hypertension** | 159 (50.3) |
| **Diabetes** | 44 (13.9) |
| **Hypercholesterolemia** | 145 (45.9) |
| **Current smoking** | 6 (1.9) |
| **Family history of premature CAD** | 24 (7.6) |
| **Prior CAD history** | 44 (13.9) |
| Mean ± standard deviation or number (%) |  |

**Table 2** compares the LV mass values obtained from MRI, contrast CTA, and AI from non-contrast CT. **Figures 1 and 2** display correlation plots and Bland-Altman analysis among the three imaging modalities.

**Figure 1.**
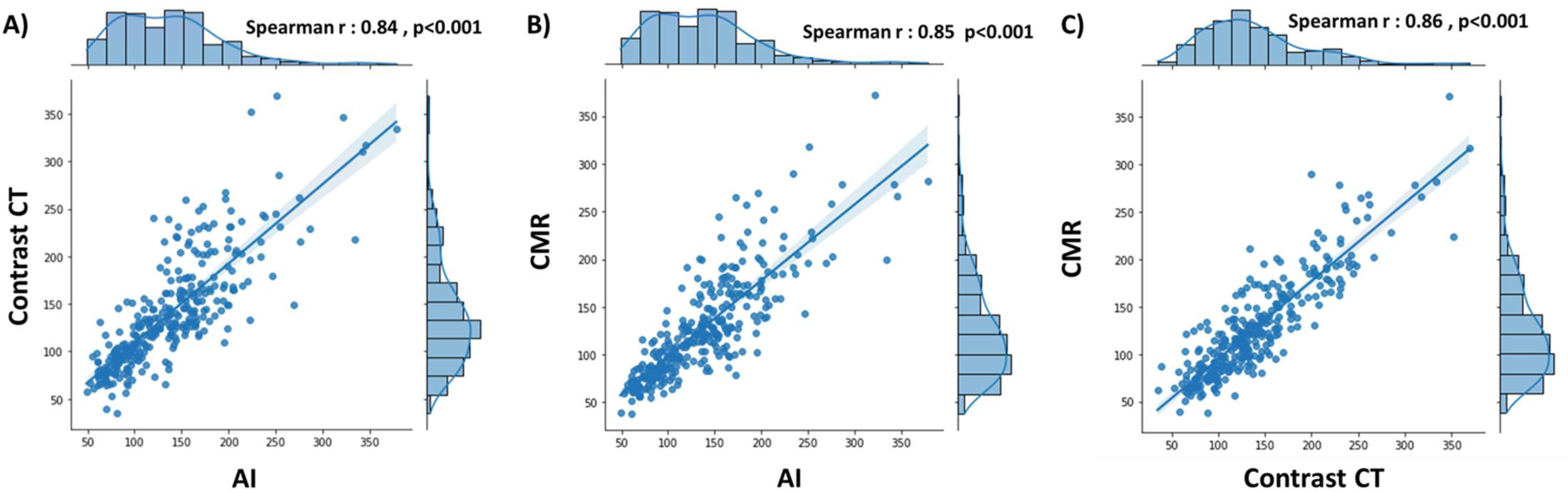
Correlation between MRI, contrast CT and AI. Blue histograms show the distributions of LV mass values in each modality. A) Contrast CT and AI; B) MRI and AI; C) MRI and contrast CT

**Figure 2.**
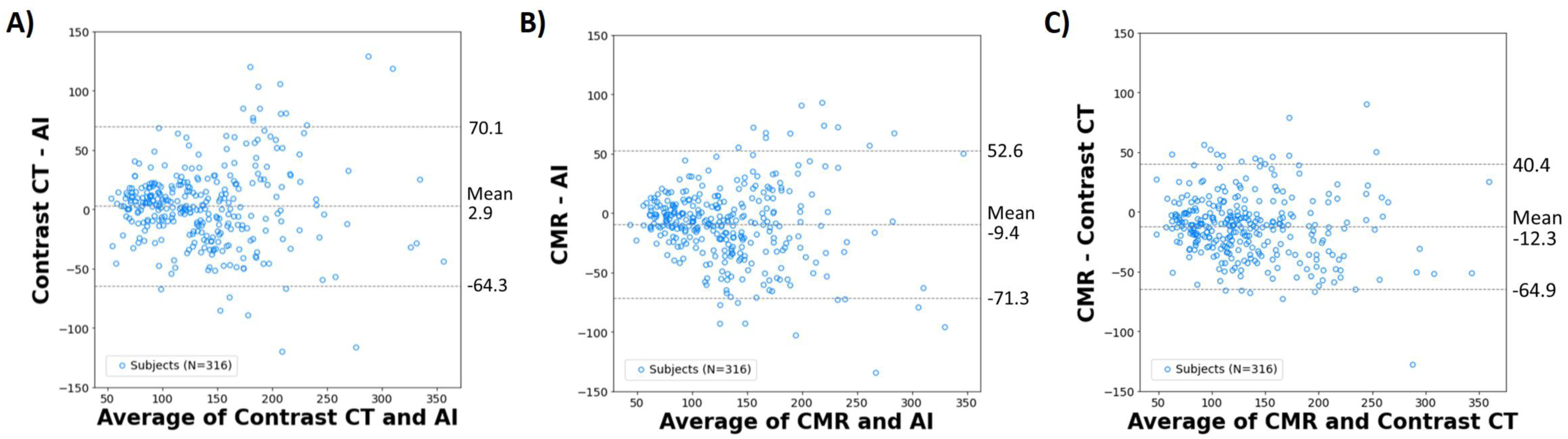
Bland-Altman analysis showing mean bias (with ±1.96 standard deviation) between MRI, contrast CT and AI. A) Contrast CT and AI; B) MRI and AI; C) MRI and contrast

**Table 2.**
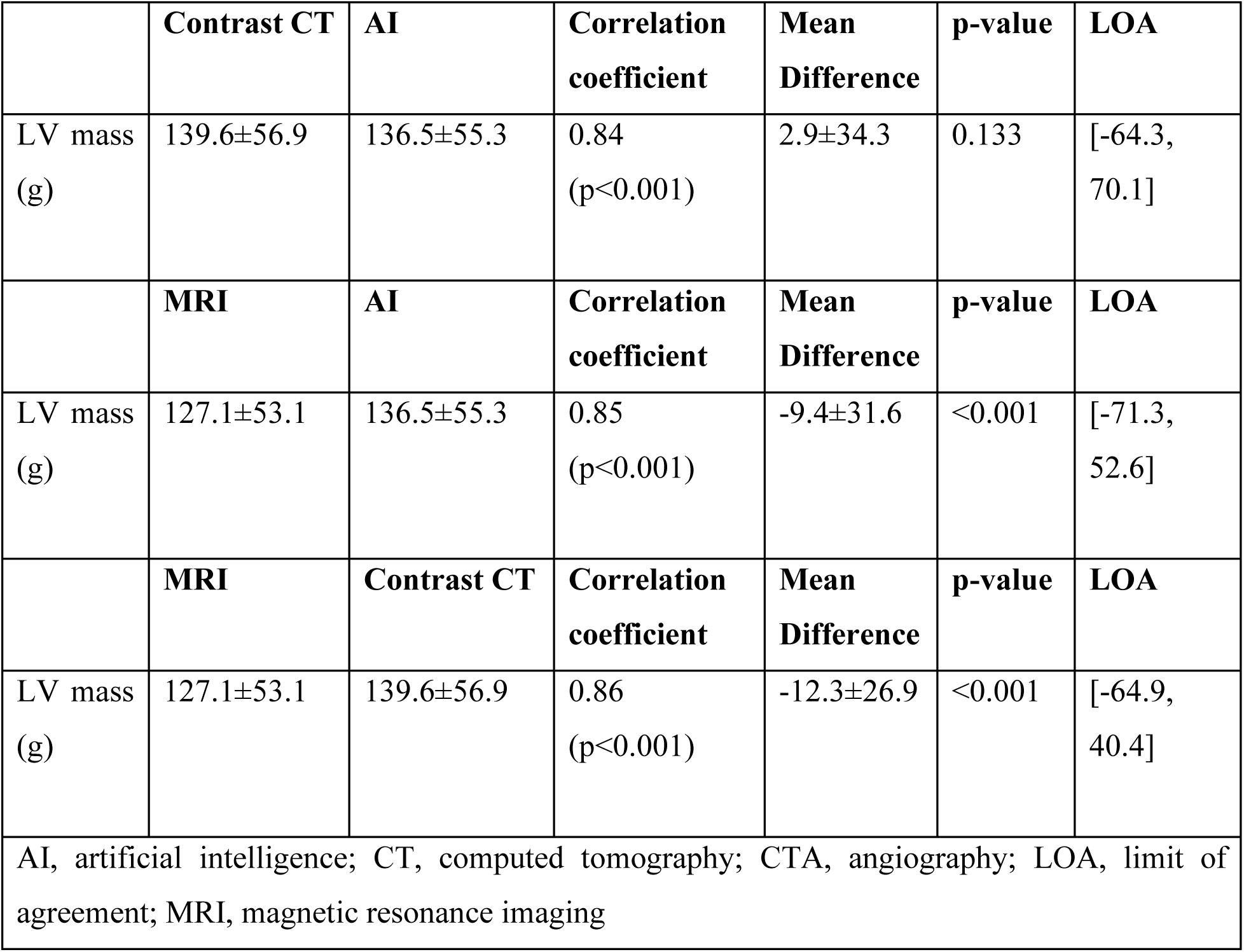
Comparison of LV mass from MRI, contrast CT and non-contrast CT (AI)

### CTA vs. AI

There was no significant difference in LV mass measures between the AI and contrast CTA (136.5±55.3 vs. 139.6±56.9, p=0.133). Further, an excellent correlation was observed between AI and contrast CTA LV mass measures (r=0.84, p<0.001) (**Figure 1**). Bland-Altman analysis showed a minimal bias of 2.9 with 95% limits of agreement between the methods of 70.1 and −64.3 (**Figure 2**).

### MRI vs. AI

When compared to MRI, measured LV mass was higher with AI (136±55.3 vs. 127.1±53.1 grams, p<0.001). There was an excellent correlation between AI and MRI myocardial mass (r=0.85, p<0.001) (**Figure 1**). Bland-Altman analysis indicated a small bias (−9.4) toward higher myocardial mass with AI, with 95% limits of agreement of 52.6 and −71.3 (**Figure 2**).

### MRI vs. Contrast CT and AI

The correlation between MRI and contrast CT was 0.86, comparable with the correlation coefficient between MRI and AI (r=0.85, p for difference=NS). Bland-Altman analysis also illustrated a similar degree of biases existed in LV mass between MRI vs. Contrast CT and MRI vs. AI (**Figure 2**).

### Subgroup analysis

The LV mass values from MRI, contrast CTA, and AI were compared separately for men and women, as shown in **Table 3**. The correlations between LV mass from contrast CTA and AI were similar for both men and women, with the correlation coefficient of 0.75 and 0.77. The mean differences were −0.9 for men and 7.8 for women. In both sexes, the LV mass values from contrast CTA and AI were found to be higher compared to MRI (**Table 3** both p<0.001). When we confined the analysis to patients who underwent CTA and MRI scans specifically for cardiomyopathy evaluation (**Table 4**), the same trend was observed. The correlation coefficient between contrast CTA and AI was 0.81, with a minimal bias of 4.4. **Figure 3** illustrates example cases with MRI, CTA, and LV endocardial and epicardial borders generated from a non-contrast CAC scan using the AI model.

**Figure 3.**
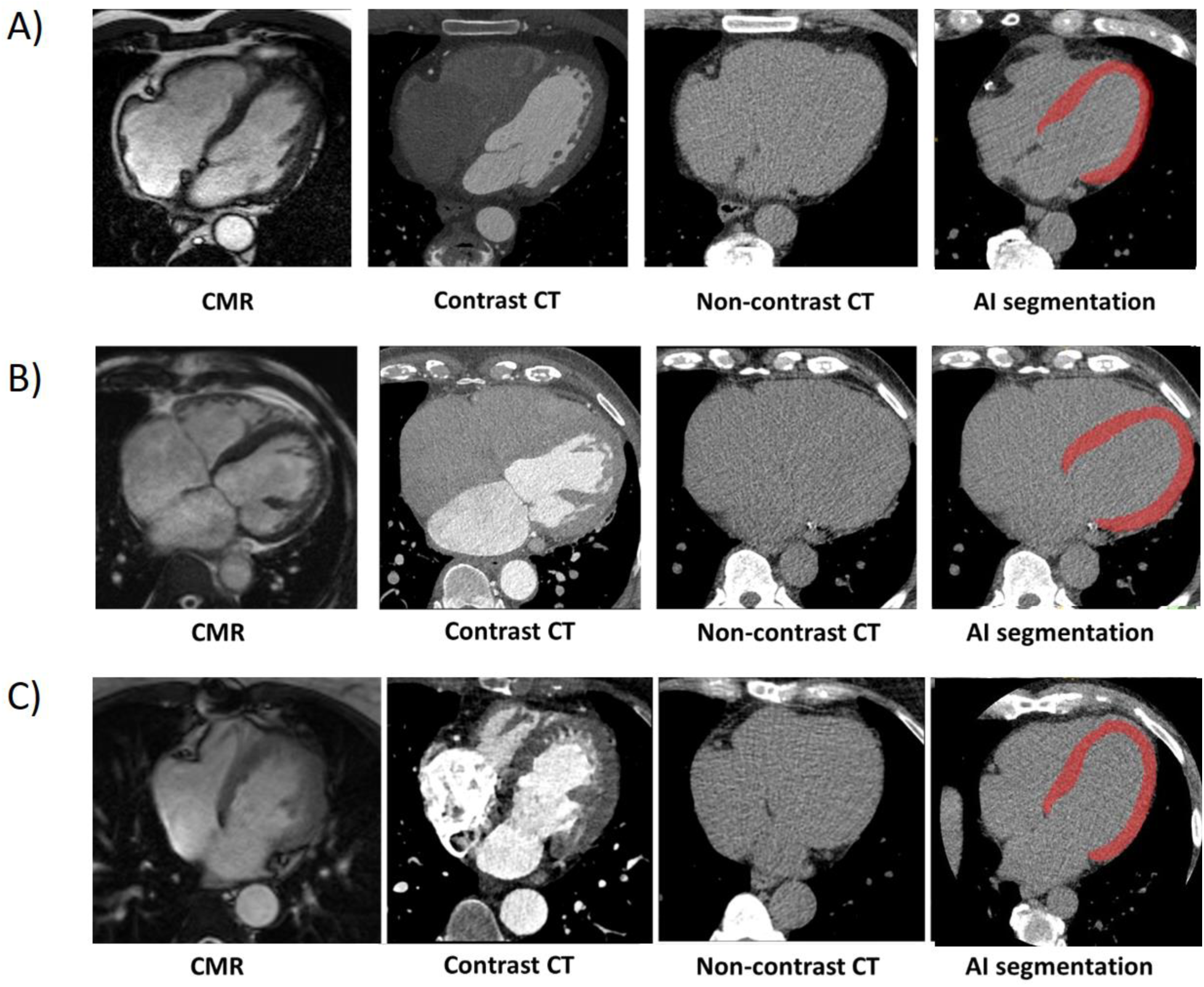
Case examples. A. Male, in their 60s (65-69), presenting with non-cardiac chest pain. LV mass by MRI: 130g, contrast CT: 149g, AI: 164g B. Male, in their 70s (70-74), presenting with shortness of breath and with a prior history of coronary revascularization, evaluation for ischemic cardiomyopathy. LV mass by MRI: 205g; contrast CT: 242g and AI: 239g C. Female, in their 70s (70-74), presenting with shortness of breath and with a history of aortic regurgitation, status post aortic valve surgery 12 years ago. LV mass by MRI: 91g; contrast CT: 104g and AI: 92g

**Table 3.** Subgroup analysis: Men and women.

|  |  |  |  |  |  |  |
| --- | --- | --- | --- | --- | --- | --- |
| Men (n=177) |  |  |  |  |  |  |
|  | **Contrast CT** | **AI** | **Correlation coefficient** | **Mean Difference** | **p-value** | **LOA** |
| LV mass (g) | 161.0±55.4 | 161.9±51.6 | 0.75 | -0.9±38.2 | 0.743 | [-75.7, 73.9] |
|  | **MRI** | **AI** | **Correlation coefficient** | **Mean Difference** | **p-value** | **LOA** |
| LV mass (g) | 148.9±51.0 | 161.9±51.6 | 0.76 | -13.1±35.6 | <0.001 | [-82.9, 56.6] |
|  | **MRI** | **Contrast CTA** | **Correlation coefficient** | **Mean Difference** | **p-value** | **LOA** |
| LV mass (g) | 148.9±51.0 | 161.0±55.4 | 0.85 | -12.1±29.1 | <0.001 | [-69.2, 44.9] |
| Women (n=139) |  |  |  |  |  |  |
|  | **Contrast CT** | **AI** | **Correlation coefficient** | **Mean Difference** | **p-value** | **LOA** |
| LV mass (g) | 111.9±46.1 | 104.1±41.2 | 0.77 | 7.8±28.1 | 0.001 | [-47.1, 62.7] |
|  | **MRI** | **AI** | **Correlation coefficient** | **Mean Difference** | **p-value** | **LOA** |
| LV mass (g) | 99.4±41.6 | 104.1±41.2 | 0.78 | -4.6±25.1 | <0.001 | [-53.6, 44.3] |
|  | **MRI** | **Contrast CT** | **Correlation coefficient** | **Mean Difference** | **p-value** | **LOA** |
| LV mass (g) | 99.4±41.6 | 111.9±46.1 | 0.76 | -12.4±23.8 | <0.001 | [-59.0, 34.1] |
| AI, artificial intelligence; CT, computed tomography; CTA, angiography; LOA, limit of agreement; MRI, magnetic resonance imaging |  |  |  |  |  |  |

**Table 4.** Subgroup analysis: patients who referred for cardiomyopathy evaluation (n=139)

| Cardiomyopathy evaluation (n=139) |  |  |  |  |  |  |
| --- | --- | --- | --- | --- | --- | --- |
|  | **Contrast CTA** | **AI** | **Correlation coefficient** | **Mean Difference** | **p-value** | **LOA** |
| LV mass (g) | 158.1±64.3 | 153.7±61.7 | 0.81 | 4.4±39.6 | 0.194 | [-73.2, 81.9] |
|  | **MRI** | **AI** | **Correlation coefficient** | **Mean Difference** | **p-value** | **LOA** |
| LV mass (g) | 144.4±58.7 | 153.7±61.7 | 0.81 | -9.3±37.4 | 0.004 | [-82.6, 63.9] |
|  | **MRI** | **Contrast CT** | **Correlation coefficient** | **Mean Difference** | **p-value** | **LOA** |
| LV mass (g) | 144.4±58.7 | 158.1±64.3 | 0.89 | -13.7±28.3 | <0.001 | [-69.2, 41.8] |

AI, artificial intelligence; CT, computed tomography; CTA, angiography; LOA, limit of agreement; MRI, magnetic resonance imaging

## Discussion

In the current study, we evaluated an AI-based method for quantifying LV mass using non-contrast CAC scans. The AI model performed fully automated LV segmentation, with a processing time averaging 22 seconds per case. Our findings demonstrated an excellent correlation and agreement of LV mass between the AI method using non-contrast CT and manual quantification from the contrast CTA. Moreover, when compared to MRI-derived LV mass, there were no significant differences in the correlation and bias between contrast CTA vs. MRI and AI vs. MRI. Similar correlations and agreements were observed between women and men, and in clinically important subgroup of patients who were referred for cardiomyopathy evaluation. To the best of our knowledge, this study is the first to comprehensively validate a fully automated AI-based LV mass evaluation from non-contrast CT by MRI, which is considered the gold standard for LV mass assessment, as well as CTA.

A few recent studies have shown the application of AI algorithms for cardiac chamber segmentation from non-contrast CT ^20,21^. However, these studies had limited small sample size and lacked a comparison with the reference standard. For example, Bruns et al. developed deep learning algorithm for chamber quantification using dual energy CT scan data from 18 patients ^21^. The algorithm was trained using virtual non-contrast images derived from dual-energy CT data and tested on standard non-contrast CT scans. Although the AI algorithm exhibited reasonable accuracy for segmentation, the quantified chamber volumes and LV mass were lower than those obtained from contrast CT scans. In our study, we utilized TS, an nn-Unet AI model, which was trained on a subset of 1204 patients with expert labels transferred from contrast scans onto aligned non contrast scans ^16^. We employed this model to quantify LV mass on ECG-gated non-contrast CAC scans in a relatively large sample size of 316 patients who underwent both coronary CTA and MRI—a gold standard for the clinical evaluation of LV mass. Our findings demonstrated a strong correlation and minimal bias in LV mass quantification between non-contrast CT and both contrast CTA and MRI scans. The AI model was trained on a wide range of different CT images (different scanners, institutions, protocols); leading to accurate LV segmentation compared to contrast-enhanced CT. Importantly, the AI-generated LV mass showed equivalent correlation and biases of LV mass between contrast CTA vs. MRI and AI vs. MRI.

Importantly in our analysis, the correlations and agreements of AI derived LV mass from non-contrast CT remained high in clinically important subgroup of patients who were referred for cardiomyopathy evaluation. It is well known that there are distinct differences in LV mass between men and women ^22^. Our subgroup analysis based on sex demonstrated a comparable correlation and agreement of LV mass in both sexes. These subgroup findings suggest the potential of the rapid and fully automated AI method can accurately assess LV mass from simple non contrast chest CT scan, regardless of sex and provide valuable information in improving the detection of adverse LV remodeling resulting from cardiomyopathies.

An important aspect of LV mass evaluation is its relatively consistent quantification across cardiac cycles ^23^. Therefore, unlike cardiac chamber volumes, which heavily depend on the acquisition cardiac phase, LV mass evaluation can be widely applicable to patients undergoing a broad range of CT scans, which include the heart within the field of view and are acquired in just one phase, such as ECG gated CAC scans.

### Limitations

Our study has several limitations. The current study was conducted at a single center utilizing specific CT and MRI scanners, which could limit the generalizability of our findings ^16^. Our evaluation of the AI model was only with ECG gated, non-contrast CT scans for CAC scanning. Further studies need to validate the performance of the AI model on non-ECG gated CT scans. Although our study has a relatively large sample size in this type of studies, the limited number of participants and selection bias from including patients who underwent CTA and MRI precludes us from establishing normal reference values. Future work including a cohort of patients with normal LV mass on CTA could be used to establish normal ranges for AI-based LV mass, which could potentially offer clinically valuable information.

## Conclusions

The AI-based automated estimation of LV mass from non-contrast CT demonstrated excellent correlations and minimal biases when compared to measurements obtained from contrast CTA and MRI. This fully automated measure could be applied for routine evaluation of LV mass in patients who have undergone a non-contrast CT scan, providing an additional imaging biomarker previously thought not measurable by this modality.

## AUTHOR CONTRIBUTIONS

DH participated in study design, data analysis, data acquisition, manuscript drafting and revisions. AS participated in study design, data analysis, manuscript drafting and revisions. RM participated in study design, data analysis, and critical revision of the manuscript. NK participated in data acquisition, data analysis, and critical revision of the manuscript. PW participated in data acquisition, data analysis, and critical revision of the manuscript. VB participated in data acquisition, and critical revision of the manuscript. DB participated in study design and critical revision of the manuscript. DN participated in study design and critical revision of the manuscript. DD participated in study design and critical revision of the manuscript. PS participated in funding acquisition, study design, data analysis and critical revision of the manuscript. All authors read and approved the final manuscript.

## ACKNOWLEDGEMENTS

This research was supported in part by grant R35HL161195 from the National Heart, Lung, and Blood Institute/ National Institutes of Health (NHLBI/NIH) (PI: Piotr Slomka), grant R01EB034586 from the National Institute of Biomedical Imaging and Bioengineering (NIBIB), and a grant from the Dr. Miriam and Sheldon G. Adelson Medical Research Foundation. The content is solely the responsibility of the authors and does not necessarily represent the official views of the National Institutes of Health.

## DISCLOSURES

D.B and P.S participate in software royalties for QPS software at Cedars-Sinai Medical Center.

D.B is a consultant for GE Healthcare. P.S has received research grant support from Siemens Medical System and consulting fees from Synektik, SA. R.M has received research support and consulting fees from Pfizer. D.N is supported by the British Heart Foundation, is a recipient of a Wellcome Trust Senior Investigator Award (WT103782AIA) and has received honoraria for consultancy and lectures from AstraZeneca. The remaining authors have no relevant disclosures.

## DATA AVAILABILITY STATEMENT

Deidentified results supporting this study are available for research purposes upon reasonable written request to the corresponding author. Access to such data is available from the date of publication and requires a Data Access Agreement, which is examined and approved by the ethics committees who approved this research.

## CODE AVAILABILITY STATEMENT

The source code can be shared using a Creative Commons NC-ND 4.0 international license upon reasonable written request to the corresponding author and requires a data use agreement.

## Notes

### Author Declarations

IRB of Cedars-Sinai Medical Center gave ethical approval for this work.

